# Differential response to cytotoxic therapy explains treatment dynamics of AML patients: insights from a mathematical modelling approach

**DOI:** 10.1101/2020.02.12.20021915

**Authors:** H. Hoffmann, C. Thiede, I. Glauche, M. Bornhaeuser, I. Roeder

## Abstract

Disease response and durability of remission are very heterogeneous in patients with acute myeloid leukaemia (AML) patients. There is increasing evidence that the individual risk of early relapse can be predicted based on the initial treatment response. However, it is unclear how such a correlation is linked to functional aspects of AML progression and treatment. We suggest a mathematical model in which leukaemia-initiating cells and normal/healthy hematopoietic stem and progenitor cells reversibly change between an active state characterized by proliferation and chemosensitivity and a quiescent state, in which the cells do not divide, but are also insensitive to chemotherapy. Applying this model to 275 molecular time courses of NPM1-mutated patients, we conclude that the differential chemosensitivity of the leukaemia-initiating cells together with the cells’ intrinsic proliferative capacity is sufficient to reproduce both, early relapse as well as long-lasting remission. We can, furthermore, show that the model parameters associated with individual chemosensitivity and proliferative advantage of the leukemic cells are closely linked to the patients’ time to relapse. They can, therefore, be used as a measure of the aggressiveness of the disease. Early assessment of these measures and incorporation into risk stratification schemes will improve risk assessment and individual treatment in AML.

## I. Introduction

**A**cute myeloid leukaemia (AML) describes a group of malignant stem cell disorders, in which functional blood cells are rapidly replaced by malignant blasts. Due to the acquisition of multiple genetic and epigenetic aberrations in hematopoietic stem and progenitor cells, those cells lose their ability to finally differentiate, which is frequently combined with an increase of their proliferative potential. These changes induce a competitive advantage and the malignant cells ultimately outcompete normal haematopoiesis. Clinically, the expansion of the malignant cells results in an acute and, if not immediately treated, fatal hematopoietic insufficiency. Whole-exome sequencing of large numbers of AML samples showed an extensive mutational heterogeneity between different patients with an average of 5 mutations in genes that are recurrently mutated [1]. The patient-specific mutational profile combined with other cytogenetic measurements at the time of diagnosis is used to categorize the patients into different risk-groups according to the European Leukemia Net (ELN) recommendations [2]. This categorization supports decisions about suitable treatment strategies.

Although treatment options have improved over the last decades, the prognosis for AML patients is still unsatisfactory. While 35 to 40 % of patients under the age of 60 are cured, older patients have a significantly worse prognosis with less than 10 % surviving after 5 years [3]. Whereas allogeneic hematopoietic cell transplantation is often the only curative approach, this option is not available to all patients and conveys its own specific risks, such as graft-versus-host disease. Therefore, the standard primary treatment for AML still consists of intensive chemotherapy including an induction therapy followed by a period of consolidation therapy. Chemotherapies are administered in a cyclic manner with usually 7 days of treatment followed by a treatment free interval. Induction and consolidation therapy include about 5 cycles of treatment. Cytarabine combined with an antracyclin are the most frequently used chemotherapeutic drugs in AML. These chemotherapies are highly cytotoxic, especially when applied in high-doses. In particular, they act unspecifically, i.e. not only affecting the malignant but also all other dividing cells, therefore, often leading to severe side effects. Even after successful primary therapy about half of the patients relapse. Those relapses represent a major challenge in AML treatment as they are both, hard to predict and difficult to treat. Therefore, the time point of relapse is a critical measure reflecting the severity of the subsequent disease course.

In a recent study, we could show that the early molecular disease dynamics are closely linked to the long-term outcome in *NPM1*-mutated (*NPM1*-mut) AML patients [4]. Therefore, we argue that it is beneficial to incorporate measures about the individual disease dynamics in the process of clinical decision-making in order to further improve personalized treatment adaptation. Technically, the assessment of molecular disease dynamics is realized by monitoring the leukemic burden in AML patients during and after therapy. However, the quantitative assessment of leukemic markers is challenging as the abundance of those mutations does not necessarily correlate with the disease load. Some mutations in genes, such as in *DNMT3A* and *TET2* are known as preleukemic mutations that can persist during complete remission [5], while others are germ line associated, such as *RUNX1* and *GATA2* [6]. Mutations in genes like *FLT3-ITD* and *NRAS* are frequently lost upon relapse. Stable lesions recommended by the ELN for measurable residual disease (MRD) monitoring are the *NPM1* mutations and gene fusions such as *RUNX1-RUNX1T1, CBFB-MYH11*, and *PML-RARA* [7]. As the measurement of mutated *NPM1* is now included in clinical routine, it is possible to track the molecular disease dynamics for these patients with high accuracy.

To gain further insights in the molecular mechanisms and individual differences in disease dynamics, mathematical models are a powerful tool, as was already shown for several diseases, such as cancer in general [8], chronic myeloid leukaemia (CML) [9, 10, 11], Alzheimer’s disease [12, 13] and Parkinson’s disease [14, 15]. Existing models of AML either address properties of the leukemic cells [16, 17, 18] or focus on the mechanisms of the disease in general [19, 20, 21]. However, so far none of these models has been specifically tailored to account for individual patients’ molecular disease dynamics and to correlate these with achievement of remission or relapse. Such a model would allow to quantitatively study the course of disease and factors influencing it *in silico* and to facilitate predictions on the relapse risk.

Earlier studies demonstrated that a mathematical model of hematopoietic stem cell (HSC) organization, which assumes a reversible switch between two functional states of HSC (i.e. a proliferative, active and a quiescent state), is able to consistently reproduce a number of experimental and clinical phenomena [22, 23, 24]. Translating this concept into the leukaemia context proved to be predictive, especially for the case of CML. Here, the theoretical analysis of the interaction between the cell cycle status of leukemic cells and the effects of tyrosine kinase inhibitors allowed to quantitatively predict individual long-term disease dynamics [25] as well as the potential for TKI dose de-escalation and, therefore, side-effect reduction [11]. Based on these insights, we here apply the same concept to capture the treatment dynamics of AML. In particular, we investigate the question, whether the leukemic reduction during chemotherapy can be mimicked without directly integrating any assumptions of a differential cytotoxic effect on leukemic cells.

In the present study we intentionally use a simple two-compartment approach to mathematically model disease and treatment dynamics in AML. We explicitly consider a chemotherapy effect operating on all actively cycling cells, independent of their mutation status. It is our main objective to test whether such a simple model description, i.e. considering an unspecific therapeutic cell kill together with an increased cell cycle activity of malignant cells, is sufficient to consistently describe the individual patient dynamics as observed in a large cohort of patients. To do so, we fit the model individually to each molecular time course of n=275 *NPM1*-mut AML patients. We further investigate the ability of the model to estimate patients relapse times as a measure of the severity of disease, as well as to improve risk stratification and predict relapse.

## II. Methods

### i. Patient Data

We used qPCR *NPM1* time courses of n=275 patients from the AML Registry of the University Hospital Dresden Carl Gustav Carus and two phase 3 trials of the Study Alliance Leukemia (SAL), namely AML2003 (NCT00180102) and AML60+ (NCT00180167). All patients gave written informed consent to participate in the study in accordance with the Declaration of Helsinki. Bone marrow aspirates were taken at irregular time points, usually at diagnosis and during primary therapy, with additional measurements during follow-up and at relapse. NPM1 quantification was processed as described in [26]. As there were two different platforms used for PCR-quantification (i.e., Lightcycler480 and Taqman 7500, both with 5’-nuclease assays) a correction factor between the two values was estimated and applied to ensure comparability between measurements (for details see Supplementary Material). In total, 69 patients presented with a molecular relapse and 57 patients did not reach remission. The median age was 53 years (range: 20 - 79). The median number of *NPM1* measurements per patients was 5 (range: 3 - 21). The medium number of therapy cycles received was 4 (range: 1 – 10). Measurements below the detection limit of 0.001 % *NPM1/ABL* (i.e. the lower limit of detection of the method) were handled as 0.001 %. All measurements after allogeneic stem cell transplantation were removed from the data set, as this treatment has a major effect on the disease dynamics and is not part of this analysis.

Patients were only included in our analysis of the accuracy of relapse time estimation if they did not reach remission at all, reached remission and relapsed within two years, or presented with later measurements confirming sustained remission during the entire two years.

We recently suggested different quantitative characteristics to describe a patients’ molecular time course (in [4]). These are: the elimination slope (*α*), the minimal *NPM1/ABL* level after primary treatment (induction + consolidation) within 9 months after treatment start (*n*), the maximum slope during relapse phase (*β*) and the time of molecular relapse (*d*) (for explanation see Figure 3A). We defined molecular relapse as the approximated time point when the *NPM1/ABL* ratio exceeds the relapse threshold of 1 % (following the suggestion in [26]). This time was approximated using a linear interpolation between the last point below the threshold and the first one above.

**Figure 1:**
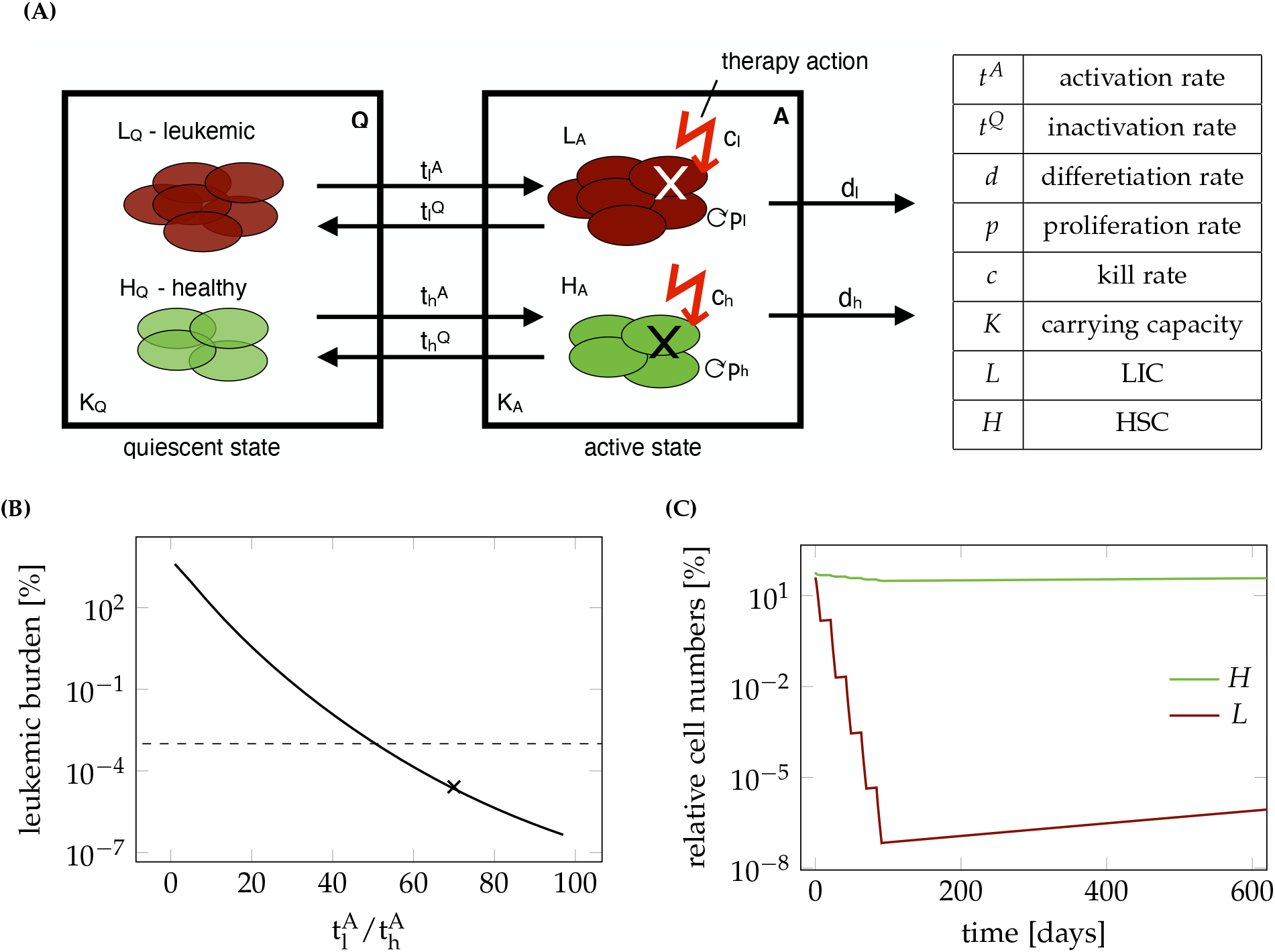
**A** Schematic overview of the mathematical model describing the dynamics of leukaemia-initiating cells and healthy stem cells in the bone marrow of an AML patient. **B** Leukemic burden after therapy depending on the ratio of leukemic activation to healthy activation 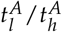. Dashed line shows detection limit for clinically measured leukemic burden. For **x** the time course is shown in Figure 1C. **C** Time course of leukemic and healthy cell numbers relative to the numbers at diagnosis with a 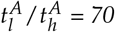.

**Figure 2:**
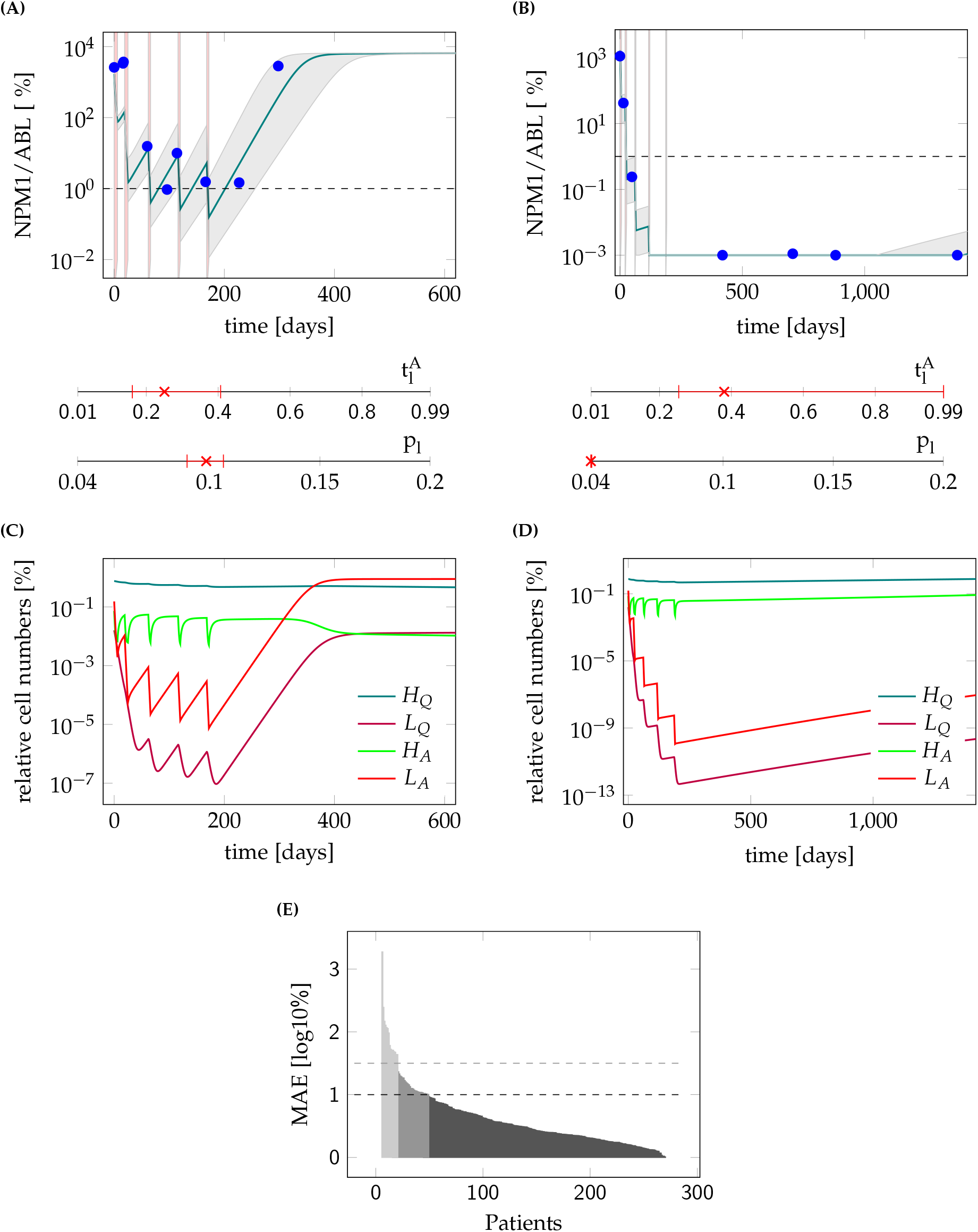
**A** Model fit to the data (PatientID = 104) and 95%-confidence interval of an example patient suffering a relapse. Mean absolute error (MAE) = 0.6 log10% **B** Model fit to the data (PatientID =3137) and 95%-confidence interval of an example patient staying in long time remission. MAE = 0.04 log10%. The red shaded regions are the times of treatment. The red marks indicate the fitted parameter values for the leukemic activation 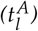 and the leukemic proliferation (p_l_) with their corresponding 95% confidence interval. **C** Time courses of cell numbers relative to the total cell numbers at diagnosis of model fit to patient from plot (A). **D** Time courses of cell numbers relative to the total cell numbers at diagnosis of model fit to patient from plot (B). H_Q_ - quiescent HSCs, L_Q_ - quiescent LSCs, H_A_ - active HSCs, L_A_ - active LSCs. **E** Mean absolute error (MAE) in log10% for all patient fits as a measure of goodness of fit. Black dashed line indicates threshold for quantitatively good fitting patients. Gray dashed line indicates threshold for qualitatively good fitting patients. For all patients with higher MAE the fit was poor.

**Figure 3:**
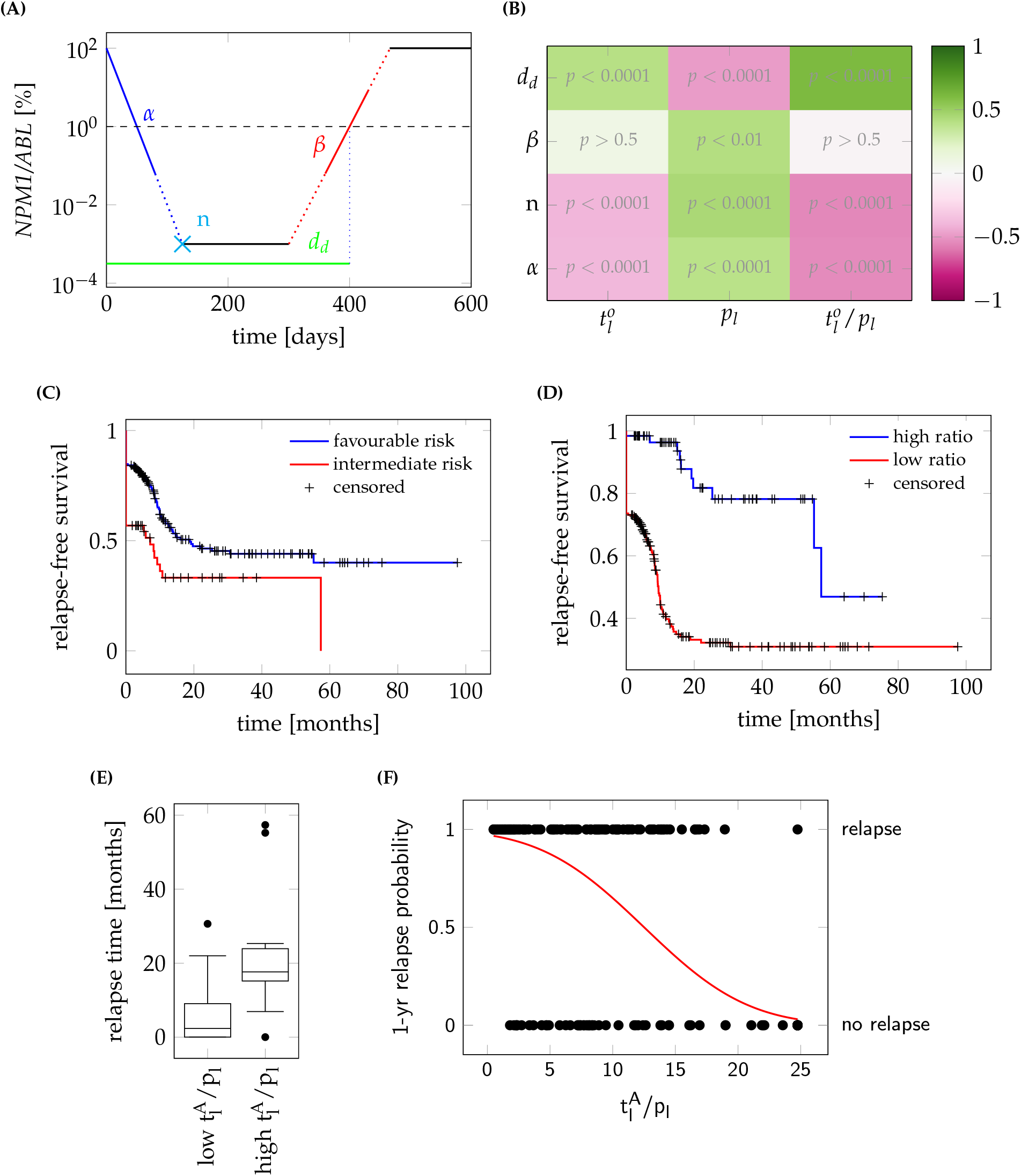
**A** Schematic overview of an NPM1 time course. α: elimination slope during primary therapy; n: minimal NPM1 level after primary treatment within 9 months after treatment start; d: time until molecular relapse; **B** Spearman correlation coefficients between the fitted parameters (i.e. leukemic activation 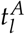, leukemic proliferation p_l_ and their ratio) and the patients molecular time course characteristics with adjusted p-values. **C** Relapse-free survival for intermediate and favorable ELN risk groups. P < 0.01 (logrank-test). HR = 1.86; **D** Relapse-free survival for high and low ratio of leukemic activation and leukemic proliferation 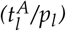 as estimated by the mathematical model with a threshold between high and low of 18. P < 0.0001 (logrank-test). HR = 5.47. **E** Boxplot for the comparison of the molecular relapse times of patients with high (n=116) or low (n=10) ratio of leukemic activation and leukemic proliferation 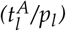 as estimated by the mathematical model with a threshold between high and low of 18. P < 0.0001 (U-test) **F** 1-year relapse probability (sigmoid function) as estimated by a logistic regression. Corresponding patient data (•) indicate the 1-year relapse status (relapse/no relapse) depending on the ratio of the two fitted parameters 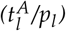.

The whole dataset used for the present analyses is available as supplementary material (Patient-Data1.csv with all patient specific information; PatientData2.csv with *NPM1/ABL* measurements).

### ii. The mathematical model

We developed a mathematical model describing the HSC dynamics in the bone marrow of AML patients (see Figure 1A). Since AML dynamics are largely driven by a pool of leukemia-initiating cells, we restrict our model to the stem cell dynamics (c.f. [17]). Similar to the general modelling concept applied for healthy haematopoiesis and chronic myeloid leukaemia [9, 22], we consider two stem cell states with different proliferative activity, i.e. quiescent and proliferating cells. Cells in these two states are assumed to differentially respond to chemotherapy. While quiescent cells, both leukemic (*L*_*Q*_) and healthy (*H*_*Q*_), are considered insensitive to S-phase specific drugs, activated leukemic (*L*_*A*_) and healthy cells (*H*_*A*_) are targeted by those substances. Assuming a finite (stem cell) niche capacity for both states (*K*_*Q*/*A*_) [27] the transition between the states is modelled as a function of the number of cells in the target state. Additionally, assuming different transition rates between the states for leukemic 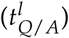 and healthy 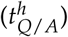 cells, the following system of ordinary differential equation (ODEs) describes the change of the number of cells (*H*_*Q*_/*L*_*Q*_) in quiescent state *Q* over time (*t*):

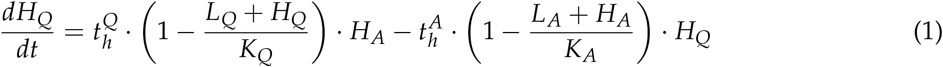

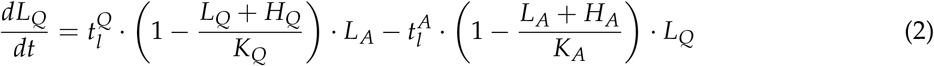

The cell numbers (*H*_*A*_/*L*_*A*_) in active state A are additionally influenced by the proliferation of the active cells with rate *p*_*h*/*l*_, and by their dependency on the total amount of cells in the current state, modelled by the carrying capacity *K*_*A*_. Furthermore, active cells can leave the stem cell state with a differentiation rate *d*_*h*/*l*_ or they can be targeted by chemotherapy with a constant kill rate *c* during treatment. The cyclic chemotherapy administration is realized in the model by a fixed chemotherapy kill rate, which can be switched on and off. These assumptions lead to the following model description in the active state *A*:

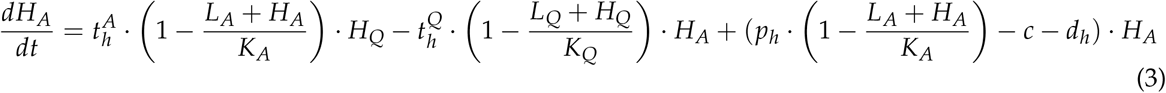

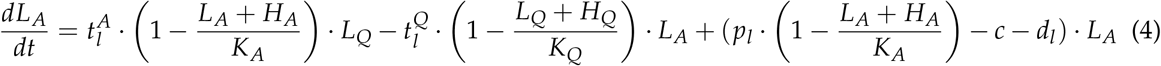

The main readout of the model to be compared with the patients *NPM1/ABL* measurements is the leukemic burden as the percentage of leukemic cells in both states combined with respect to all cells in the states:

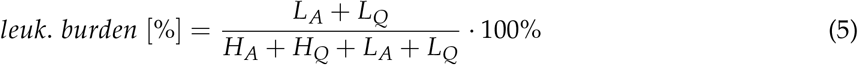

To compare model results with the data, all values below the detection limit of the qPCR method of 0.001 % were set to this limit. As it is common to have *NPM1/ABL* ratios of up to 1000 %, because of the higher abundance of the *NPM1* transcript compared to the reference gene [28, 29] the percentage of leukemic cells from the model was increased by a factor of 100 to translate them into the *NPM1/ABL* ratio and make the model results comparable with the measurements.

### ii. Model parameters

Most values for the model parameters were taken from the literature (see below) and treated as constant values for all patients. To account for patient-specificity, the aggressiveness of the leukemic clone and the individual chemosensitivity were modelled by the leukemic proliferation rate (*p*_*l*_) and the leukemic activation rate 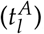, which were estimated per patient. The transition rate of leukemic cells into quiescence 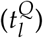 was kept constant for all patients and was set to 0.002 per day. It was obtained by fitting the model to all patients multiple times, each time with a different inactivation rate to find the globally best fitting rate. The other model parameters were set to plausible values based on available literature data:

- **Chemotherapeutic kill rate (***c*) was kept constant to ensure identifiability of the fitted parameters. It was set to 0.99 per day to account for the high total leukemic cell kill, as reported for an in vivo mouse study [30].
- **Proliferation rate of healthy cells (***p*_*h*_) was set to once every 25 days. It was estimated that healthy hematopoietic stem cells divide once every 30 days [31]. But as the actual proliferation rate in the model depends on the niche space in the current state and hence this rate would only be reached for an empty state, the rate was set to this higher value.
- **Stem cell differentiation rate (***d*_*h*/*l*_) was set to the value of the proliferation rate without influence of the capacity (1/30 per day) for both leukemic and healthy cells.
- Activation rate for healthy cells 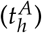 was set to 0.01 per day and the **inactivation rate** 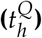 to 0.2 per day, based on the functions for these values from the original model [9].
- **Carrying capacities for each state (***K*_*A*/*H*_) were based on the value of 105 cells as previously described [9], which is about a factor 10 smaller than actual stem cell numbers in a patient’s bone marrow.

The competitive advantage of leukemic cells compared to healthy cells originates from their increased proliferative potential [32, 33], which is represented in the model as an increased leukemic proliferation rate of up to once every 5 days.

When fitting the model to patient data, the leukemic activation rate 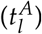 and the leukemic proliferation rate (*p*_*l*_) were estimated by minimizing the weighted log-likelihood, where negative *NPM1/ABL* measurements were double weighted to account for the possibility of these values being far below the detection limit. Differences between values were calculated on the log10 scale. For all patients, 100 random starting points were generated for fitting to eliminate the dependence on the start value and to increase the probability of reaching the global minimum. In order to avoid overfitting, we assessed the identifiability of the fitted parameters using the profile likelihood (see [34] for details). In the corresponding likelihood landscape in Supplementary Figure S1 it is shown that the identifiability is given for both free parameters.

For an additional, easier interpretable measure of goodness of fit the Mean Absolute Error (MAE) was estimated by computing the mean divergence of the model from the measurements for each patient fit on the log10 scale.

### iv. Statistical analysis

General bivariate correlations were quantified using the Spearman rank correlation coefficient (*ρ*). P-values for multiple pairwise correlations were adjusted using Bonferroni correction. To specifically measure the agreement between relapse times estimated with the model and estimated from the clinical data we applied the Lin’s concordance correlation coefficient (*ρ*_*c*_). The (Mann-Whitney) U-test was applied to compare rank distributions (i.e., shift in the medians) of two independent groups of patients. The Kaplan-Meier method was used for relapse-free survival analysis, using the time until recurrence/death. The logrank-test was applied to compare the survival of two independent groups. A cox regression was conducted for hazard ratio (HR) estimation. The threshold of the parameter ratio to discriminate between the two groups was determined by maximizing the HR with the additional requirement of at least 10 target events (i.e. relapse) in each group. Odds ratios were determined by logistic regression. Identifiability of the model parameters was assessed using the profile likelihood approach, as described in detail in [34]. The likelihood landscape is derived by estimating the profile likelihood for all parameter combination. The 95%-confidence intervals for the model parameters were derived following the likelihood-based confidence interval definition by Meeker and Escobar [35].

All analyses were done with MATLAB R2018b (The MathWorks, Inc., 1994-2018).

## III. Results

### i. Reduction of leukemic burden can be modelled without assuming a different chemotherapeutic kill effect on normal and leukemic cells

In order to reproduce the molecular dynamics of AML we developed a mechanistic mathematical model of the competition of leukemic and healthy stem cells in the bone marrow. This modelling approach is in line with earlier works in which a minimal, two-compartment model of HSC organization was applied to describe various phenomena such as stem cell competition, aging and leukaemia [22, 9, 23, 36, 11, 37]. The model describes two functional states between which the cells can reversibly transit depending on the capacity in the target state. While in the “active state” the cells proliferate and can potentially differentiate, they stay inactive in the “quiescent state” (Figure 1A). Both healthy hematopoietic stem cells (HSCs) and leukaemia-initiating cells (LICs) can adopt each state. However, while their transition dynamics between the two states differ, leukemic and normal cells are competing for shared resources (such as limited niche space). The parameters for the healthy cells were taken from the literature as described in Materials and Methods. The routinely used chemotherapeutics D-arabinosyl cytosine (AraC) and Daunorubicin effect all dividing cells during S-phase equally [38]. Therefore, our first question to be answered was, whether it is possible to assume an equal cell kill on both active HSCs and LICs during chemotherapy and still achieve a leukemic clearance. Therefore, we defined all parameters for the leukemic cells to be the same as for the healthy ones, while only the leukemic activation rate 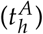 was increased systematically. Figure 1B shows the leukemic burden at the end of 5 chemotherapy cycles depending on the assumed value of leukemic activation. The figure indicates that an elimination of the measurable leukemic burden (for a detection limit at 10-3 %) can be consistently explained even without differences in the chemo-induced kill rate if an elevated activation of leukemic cells by a factor of 50 is assumed compared to healthy cells. The exemplary time course in Figure 1C confirms the rapid decline of leukemic cells during therapy, while the healthy cells stay at a high level. This adheres to the general concept that leukemic cells are almost constantly proliferating without periods of extended quiescence and reflects the typical leukemic phenotype [39]. It shows further that this model is able to reproduce a broad bandwidth of treatment induced reduction of leukemic burden (reaching from 1 to 6 log-reduction), which is also confirmed in different AML patients during chemotherapy [29].

### ii. A simple mechanistic model is able to mimic the molecular dynamics of most NPM1-mut AML patients

After having demonstrated the general ability of the model to mimic the leukemic reduction during therapy we analysed whether the model can also account for the interpatient-variability in treatment response and dynamic behaviour. For this analysis, 1567 *NPM1/ABL* measurements of 275 patients with a median number of 5 measurements per patient were available. We assessed the ability of the model to reflect a large variety of time courses, including remission (n=149), unresponsiveness to therapy (n=57) and relapse cases (n=69). Therefore, we adjusted the individual activation rate of leukemic cells 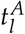, as well as the individual leukemic proliferation rate *p*_*l*_ for each patient’s time course of leukemic burden to find a patient specific parameter combination that optimally describes the individual disease dynamics, while all other parameters were kept constant. In Figure 2A and 2B there are two examples of fitted patient time courses along with the 95 % confidence interval for the optimal fit. These examples illustrate that the model reproduces the characteristic time course of AML patients presenting with either a molecular relapse or a sustained molecular remission. Although, both patients received very similar chemotherapies, it can be seen that for the relapsing patient the *NPM1* burden is not eliminated after the therapy (Figure 2A), while for the other patient (Figure 2B) the value is below the detection limit. Hence, the slightly higher leukemic activation 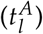 and the lower leukemic proliferation (*p*_*l*_) are sufficient to explain the difference between relapse and sustained remission. Further, we observed, that a high leukemic activation rate is linked to a fast decline of leukemic burden during therapy, an effect which is illustrated clearly by the good responding patient in Figure 2B. Therefore, we conclude that this rate estimate can be interpreted as a measure for the patient’s individual chemosensitivity. Also, we observed, that a high leukemic proliferation rate is linked to fast leukemic regrowth, which can be seen in the poorly responding patient in Figure 2A. From this we conclude that the estimated leukemic proliferation rate provides a measure of the tumour intrinsic aggressiveness.

Looking in more detail how the different cell populations within our model respond to the treatment (Figure 2C and 2D), one observes that the difference between a good and a poor therapy response is within the leukemic cell population, with the good responder showing steeper decline during therapy as well as slower regrowth afterwards. In the poor responder the number of leukemic cells in the model also drops to a vanishingly low proportion of their initial numbers during treatment. Nonetheless, a relapse occurs within a few months, because of the fast regrowth. Furthermore, a decrease of the net proliferation rate during leukemic regrowth after chemotherapy can be observed (as the curves (*L*_*A*_ and *L*_*Q*_) flatten after an exponential growth phase around 350 days after diagnosis). This observation agrees with a recent study by Akinduro et al., where the authors analysed the growth of AML cells after transplant in a mouse model [40]. The high number of quiescent HSCs is also in accordance with experimental data, as it was recently shown that in a leukemic mouse bone marrow HSCs were mostly in a quiescent state [41]. This mechanism is also referred to as “self-protection” by which HSCs avoid environmental effects in the quiescent state [42].

The quality of the individual fits was evaluated using the mean absolute error (MAE) of each model fit, which corresponds to the average residual between model fits and available data points on the log10 scale. We observed that fits with a MAE of up to 1 could quantitatively describe the patient’s course of disease. Hence, at least 227 of the 275 patients can be sufficiently described by the individual model fits (Figure 2E). Patients with an average error of up to 1.5 log could still be qualitatively described (n = 260).

Looking more closely at the 15 patients in which the model could not optimally fit to the time course data, it becomes clear that for most of them (10) the model as it is cannot account for the strong leukemic reduction found in the data (example patient in Supplementary Figure S3A). For 2 of the patients the very fast regrowth is not reproducible with the model and for 3 patients a chemo-resistance arises during treatment of a relapse, which is not yet captured by our current model (both can be seen in example patient in Supplementary Figure S3B).

Taken together, we could show that the proposed simple model is indeed able to reproduce most of the molecular behaviour of *NPM1*-mut AML patients, which can now be used to analyse the molecular characteristics of the course of disease.

### iii. The model parameters are closely linked to time course characteristics

Based on the patient individual model fits we obtain a pair of model parameters (leukemic activation rate 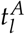 and leukemic proliferation rate *p*_*l*_) for each specific patient time course (distribution of the parameters in the cohort can be found in Supplementary Figure S2). In order to investigate how these model parameters correlate with typical, clinically relevant characteristics of the *NPM1/ABL* dynamics, we perform a systematic correlation analysis. For a quantitative description of the relevant *NPM1/ABL* time course characteristics we use a parametrization based on the elimination slope (*α*), the minimal NPM1 level after primary treatment (induction + consolidation) within 9 months after treatment start (*n*), the maximum slope during relapse phase (*β*) and the time until molecular relapse (*d*_*d*_) (see Figure 3A, [4]). Figure 3B provides a summary of pairwise comparisons between the individually fitted model parameters and the characteristics of treatment response. Generally, for all characteristics but the relapse slope *β*, a significant correlation with a correlation coefficient of up to 0.6 was found. Furthermore, the leukemic activation 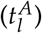 and leukemic proliferation (*p*_*l*_) are conversely correlated with all described time course characteristics. That means, that later molecular relapse is linked to increased leukemic activation, but decreased leukemic proliferation. This supports the earlier conclusion, that the leukemic activation is a measure for chemosensitivity, whereas the leukemic proliferation provides a measure for the aggressiveness of the disease. Furthermore, this anti-correlation with the time course characteristics suggests that the ratio between the two parameters 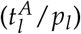 reflects a good measure for the overall severity of the patient’s individual course of disease.

From a clinical perspective it is most important to know whether and when a patient will relapse or not, and to estimate the chance for relapse-free survival. Using the approximated time point of molecular relapse as a surrogate for haematological relapse (including irresponsiveness), we studied whether a division into high and low ratio of estimated leukemic activation and leukemic proliferation (i.e. 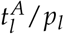; threshold at 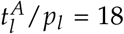, determined by maximizing the difference between both groups, details in Materials and Methods) can improve the current ELN risk stratification (stratifying into high and low risk). Figure 3C and 3D support the notion that both variables are indeed correlated with relapse-free survival. However, the differences in the Kaplan-Meier plots indicate that this effect is considerably more pronounced using the ratio of leukemic activation and proliferation. Comparing the hazard ratio (HR) between the two stratifications further supports this impression (HRratio=5.5 (95% CI: 2.8 to 10.4); HRELN=1.9 (95% CI: 1.2; 2.8)). A clear difference in the time of relapse can be seen, when directly comparing the molecular relapse times of the high and the low parameter groups (Figure 3E). Patients with a high parameter ratio 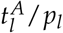 tended to relapse in median 15.3 months later. Moving away from the division into two risk groups, we apply a logistic regression approach to estimate to which extend the estimated ratio 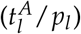 correlates with the probability to relapse within the first year after diagnosis (Figure 3F). The odds ratio of 1.17 [95% CI: 1.11 to 1.22] indicates, that an increase in the ratio by 5 units more than doubles the chance of experiencing a relapse within the first year. However, the nearly linear relationship between the ratio 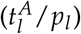 and the relapse risk indicates that the ratio 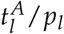 alone cannot reliably predict relapse for a particular patient, although it can be useful in combination with other measures to improve risk prediction.

### iv. Clinically relevant characteristics can be estimated using the mathematical model

In the clinical context the time of molecular relapse is a highly relevant measure. For this reason, we compared the molecular relapse time *approximated* from the data (*d*_*d*_, definition can be found in Material and Methods) with the *estimated* molecular relapse time predicted by the model (*d*_*m*_). Figure 4A shows the corresponding scatter plot for all patients for which a relapse time within 2 years could be approximated (n=175). As expected, there is a good accordance for most patients, while only for 13 out of the 175 patients (7%) the *estimated* molecular relapse time from the model diverges more than half a year from the molecular relapse time *approximated* from the data. The mean divergence for all patients is 1.9 months. However, the larger differences for some patients raises the question why the model is incapable of mimicking their course of disease.

**Figure 4:**
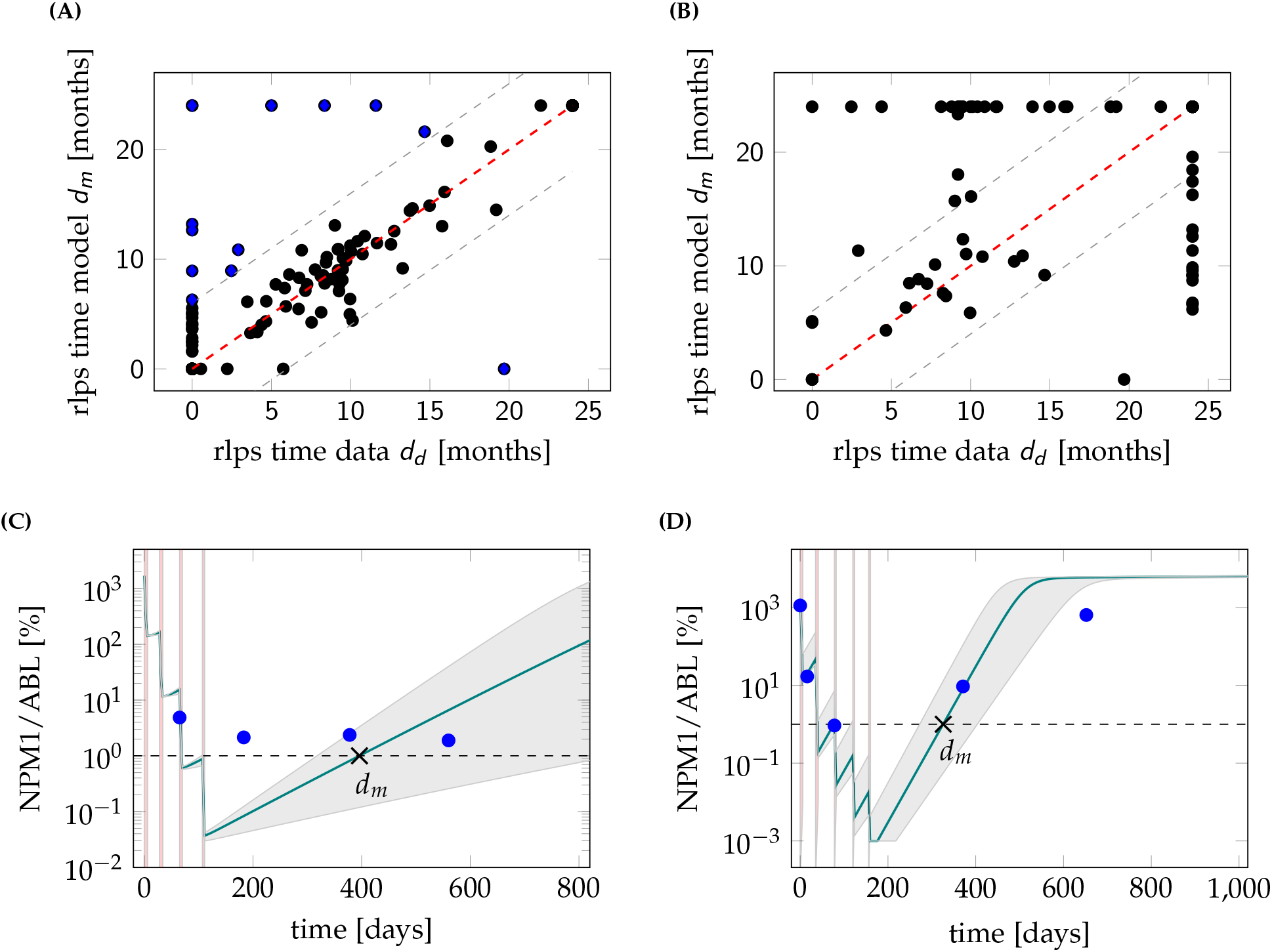
**A** Scatter plot for the comparison of the molecular relapse time approximated from the patient data d_d_ and the molecular relapse time estimated from the fitted model d_m_ for each patient. ρ_c_ = 0.90. Perfect accordance is indicated with dashed red line. Gray dashed line indicates divergence by half a year. Points with divergence larger than half a year are marked blue. **B** Scatter plot for the comparison of the molecular relapse time estimated from the fitted model to the first 9 months d_m_ of the patients and the molecular relapse time approximated from the data of these patients d_d_. ρ_c_ = 0.37. **C** Example fit (PatientID = 3751) and 95% confidence interval for a patient where the model is not able to capture the molecular course of disease. Dashed line shows remission/relapse threshold. d_d_ is 0 days (no remission reached), d_m_ is 396 days. **D** Example fit (PatientID = 3621) and 95% confidence interval for a patient where the sparseness of data points makes it impossible to reliably approximate the molecular relapse time. Dashed line shows remission/relapse threshold. d_d_ is 0 days (no remission reached), d_m_ is 326 days.

Studying the 13 critical patients more closely it appears that the model is insufficient to describe at least 6 of them for technical reasons, i.e. very fast regrowth, very high chemosensitivity or stable tumour levels, that are not depicted in the model (see example in Figure 4C). To explain a stable tumour level over a longer time (as in the example patient), an additional intrinsic leukemic suppression (such as an immunological component) would be necessary in the model. For the other 7 patients with poor accordance of the relapse times, it is the sparsity of the available measurements that limits a reliable *approximation* of the relapse times from the data. For the example patient in Figure 4D the measurements suggest that a remission could not be reached, but it is conceivable that the tumour burden decreases further during the time of therapy. Therefore, it is very likely that a temporary remission occurs at the end of therapy, as suggested by the model fit. But just from the available data both conclusions cannot be excluded. Therefore, it is possible that the *estimated* relapse time from the fit is closer to the true relapse time than the *approximation* from the data. Hence, we conclude that the model, although not able to perfectly describe every single course of disease, might indeed harbour additional information about the true course between available measurements.

### v. Predicting the relapse time based on the early time course is not reliable

We could show that the ratio 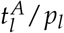 is correlated to the *approximated* time of molecular relapse and suggested that the model can help to estimate the particular time point of molecular relapse occurrence. Moving a step further, we raise the question whether an early (and clinically even more relevant) prediction of the relapse behaviour can be made with comparable precision. From a clinical perspective, 9 months after treatment start is a time point when a decision about future treatment options for a particular patient is still relevant. We chose this time point as a cut-off until which the models were fitted, given there are at least 3 data points within this time period (n=89). Figure 4B compares the relapse times *predicted* by the model based on these truncated time courses with the *approximated* relapse time from the complete data. It appears that reliable model *predictions* cannot be achieved for the majority of cases. In fact, for more than 40% of the patients the divergence is more than 6 months and the mean difference over all patients is 6.6 months. This visualization indicates that a reliable *prediction* of relapse occurrence cannot be achieved if only sparse measurements during the initial treatment response during of the first 9 months of treatment are considered. However, it cannot be excluded from our current study that additional diagnostic parameters together with close meshed short term time course data convey more information for better predictions.

## IV. Discussion

In this study we have developed and validated a simple mechanistic, mathematical model that is able to correctly reflect the individual molecular disease dynamics of *NPM1*-mut AML patients treated with cytotoxic drugs. As the model describes AML treatment response as the result of the competition between more proliferative leukemic and less proliferative healthy stem cells, we can use this model to also functionally analyse the underlying mechanisms and contribute to the understanding of AML remission and relapse. In particular, we explicitly assumed that both healthy and leukemic stem cells can be in a quiescent and chemotherapy-insensitive state, while they are equally targeted by chemotherapeutics if they are in an actively cycling state. Based on these assumptions, we addressed the question of how the individually fitted model parameters for each patient are linked to clinically relevant measures, such as the *approximated* time of molecular relapse.

The dynamics of AML pathogenesis and treatment have attracted several modelling approaches. However, few models explicitly address the competitive imbalance between leukemic and healthy haematopoiesis, and are validated with larger patient cohorts. To our knowledge, only a study by Thomas Stiehl et al. [43] follows a similar approach in which the time after induction therapy is considered. However, this model focuses only on the time after therapy, starting always with a state in which no leukemic cells are detectable. Thereby, patient specific differences in their therapy response and treatment regimen are neglected. Considering earlier clinical findings [26] as well as our patient-specific analysis, this restriction to homogeneous treatment response limits the generalizability of this modelling approach.

The overall agreement between measured patient time courses and the optimally adapted model dynamics allows us to conclude that our model captures a set of features that are characteristic for the response of AML patients to cytotoxic treatment. Most importantly, this correspondence ensures us that the general notion of hematopoietic or leukemic stem cells reversibly changing between states of differential proliferative activity and chemosensitivity, can also be applied in the context of AML. Strikingly, we did not even assume a different chemotherapeutic effect for dividing cells, irrespective of whether they are leukemic or healthy cells. This reflects the mechanism of action inherent to S-phase specific drugs which effects *all* dividing cells. From our model estimates we conclude that the unregulated and strongly increased activation of leukemic cells into cell cycle is sufficient to describe a preferential targeting of this population and to explain the typical remission behaviour. And although *NPM1*-mut AML is more chemosensitive than other AML subtypes [44], the model could be transferred, as also unresponsiveness to treatment can be captured by the model. Furthermore, our simplistic model provides additional explanations for other phenomena observed in AML, which are not explicitly included in the model setup. One of these phenomena is that the leukemic proliferation rate decreases during regrowth [40]. This can be explained with the limitation of the proliferation by the available stem cell niche capacity. During leukemic expansion the niche is rapidly filling, leading to a decreased proliferation. Another phenomenon is the high number of quiescent HSCs in AML [41], which can be explained by the fast activation of leukemic cells into the active state. Therefore, the limited niche capacity is rapidly populated by leukemic cells, pushing the healthy cells to stay quiescent. This is also in consent with findings by Miraki-Moud et al. who showed that bone marrow failure in AML is not caused by depleting HSC numbers but impairing their differentiation [45]. Another phenomenon is the increased proportion of active cells within the leukemic bulk in patients with a good therapy response compared to a poor therapy response [46] (Supplementary Figure S4). This can be explained by the link between high leukemic activation and a high chemosensitiviy. The higher leukemic activation leads to more active leukemic cells, which can be easier killed by the treatment.

Further, we could show, that the two individually fitted model parameters, namely the leukemic activation rate and the leukemic proliferation rate, are closely linked to the patients’ time course characteristics, e.g. the elimination slope, the NPM1 burden after primary treatment and the time of molecular relapse. As the leukemic proliferation is assumed to be a measure for the tumour aggressiveness, we expected to see a close link with the relapse slope *β*. However, only a weak correlation was found. The reason for this can be found in the imprecise estimation of the relapse slope on the basis of too sparse data, as the strong correlation (*ρ* = 0.97) with the relapse slope estimated from patients’ model time courses suggests. The leukemic activation rate, which is assumed to be a measure for the patient’s chemosensitivity has a less clear impact on the time course, as it has medium influence on all characteristics, except for the relapse slope. In general, we found, that both parameters combined as a ratio show the closest link to the time course characteristics. Using this ratio of leukemic activation and proliferation from the model to separate the patients into two groups (high and low estimated ratio) improves the risk stratification into favourable and intermediate risk patients compared to the ELN scheme (Figure 3D). This supports our previous claim, that the close monitoring of molecular disease markers and incorporation of the molecular disease dynamics in risk stratification schemes will improve assessment of the severity of the patients’ AML [4].

Nonetheless, the inability of the model to reproduce all patients’ time courses shows that the model lacks some further regulations that cannot be neglected for some patients. Likely candidates for such a regulation could be the immune response, as suggested by an increased activity of NK cells in patients with longer relapse free survival [47], the emergence of new leukemic clones at relapse with possibly different properties than the clone at diagnosis [48, 49], or the ability of AML blast cells to de-differentiate [50].

When using the model to predict the individual patients time of relapse based on the measurements of the first 9 months, the accuracy was very limited. One reason is the above-mentioned inability of fitting some of the patients’ time courses. The other reasons are the sparseness of the data and the measurement error, which make it difficult to get an exact fit with the model and to get an exact approximation of the patients’ molecular relapse time. A stringent MRD monitoring as proposed by Rautenberg et al. [51] would help to overcome these shortcomings in the future. We are convinced, that with such an improvement of the data, the model would be able to predict a relapse for AML patients.

In conclusion we showed in this study that the fitted parameters of a simple mathematical AML model can consistently describe individual time course characteristics in the majority of the analysed patients. Furthermore, we could show that the model can help to improve the understanding of the complex dynamics and mechanisms that interact in AML. Finally, we showed that the model can improve the classification of disease severity and the state-of-the-art risk predictions.

## Data Availability

The datasets supporting this article have been uploaded as part of the supplementary material.

## VII. Authors’ contributions

HH developed the mathematical model, carried out the statistical analyses and drafted the manuscript. IR and IG gave regular feedback during all analyses and helped draft the manuscript. CT and MB conceived and designed the study, coordinated the clinical data collection and critically reviewed the manuscript. All authors gave final approval for publication and agree to be held accountable for the work performed therein.

## VIII. Ethics statement

The study was approved by the ethical board of the Medical Faculty of the TU Dresden (EK98032010).

